# Meta-analysis of Cannabis Use Identifies Shared Genetic Loci with Sleep and Circadian Rhythms

**DOI:** 10.64898/2026.04.14.26350867

**Authors:** Jesse Valliere, Satu Strausz, Cynthia Tchio, Oona Shigeno Risse-Adams, Nasa Sinnott-Armstrong, Hanna M Ollila, Richa Saxena

## Abstract

Cannabis use is an increasingly common therapeutic for a variety of chronic diseases. In addition, people with sleep problems may self-medicate using cannabis products. However, genetic architecture of cannabis use and its shared genetic predispositions with sleep traits has not been systematically examined. We performed a meta-analysis of cannabis use within the All of Us and UK Biobank cohorts, consisting of 152,807 cases and 220,272 controls. Our meta-analysis identified 39 independent loci, including the previously reported *CADM2* locus associated with cannabis use and replicating previous work. Additionally our associations include neuronal and sleep-regulating genes such as *HTR1A, RAI1, SLC39A8*, and *NCAM1*. Moreover, tissue-specific analyses revealed that the genetic architecture of cannabis use is heavily enriched within the central nervous system and specific brain cell types. In addition, we observed significant positive genetic correlations with clinical insomnia, insomnia-related medication usage, and objectively measured nighttime physical activity, alongside negative correlations with morningness chronotype and daytime activity. Fine-mapping and colocalization analyses identified shared genetic signals between cannabis use and clinical insomnia including a near-perfect colocalization at *SLC39A8* and *CADM2*. Together, these results highlight the shared genetic risk between cannabis use and sleep disorders. Additionally, our findings indicate the importance of investigating the genetic effects of cannabis use as its use becomes more widespread, both recreationally and medicinally.

## INTRODUCTION

Cannabis is the most commonly used drug with over 219 million users estimated worldwide^1^. In 2021 the prevalence of Cannabis use was 12% in countries where it was legalized at the time, whereas in non-legalized countries estimated cannabis use prevalence is 5.4%^2^ and is expected to grow.

There are two primary uses for cannabis. It is typically used either as a recreational drug similar to alcohol or other drugs. Cannabis products are additionally prescribed as a treatment. It is used for managing pain in chronic illnesses, reducing muscle spasms in multiple sclerosis, increasing appetite in weight loss including anorexia nervosa, or for sleep problems due to its sedative and anxiety reducing properties^3 4 5^.

Importantly, public perception has morphed significantly within the past 20 years, with daily or near-daily consumption surpassing reported alcohol use in the United States in 2022 ^6^. While this shift has been seen on a cultural level within the United States, funding and research for cannabis has been more limited than other recreational drugs. Given the rising popularity of cannabis use both recreationally and in a medicinal context, it is important to observe and understand its effects on physical and mental health, diseases and associated biological factors, utilizing proxy phenotypes until more data can be obtained on chronic use.

As cannabis is relatively easily accessible in several countries, individuals also have the possibility to self-medicate. A recent study^7^ showed that individuals with sleep problems use cannabis for this purpose. However, it is possible that prolonged use of cannabis products increases tolerance and can result in increase in use and lead to cannabis use disorder. However, the exact biological mechanisms that connect cannabis use to sleep problems and vice versa are not yet fully understood.

Genetic studies provide an opportunity to understand biological pathways underlying diseases and traits. Previous studies of cannabis use have previously found eleven genetic loci for lifetime cannabis use and connected cannabis use with other recreational drugs, and with cannabis use disorder^8^. Additionally, cannabis use disorder has been extensively assessed in the context of schizophrenia and psychiatric traits, where cannabis can trigger a psychiatric episode^9^. The individual genetic variants or genetic correlations similarly reflect this association in previous work.

In this work we wanted to expand the genetic associations for cannabis use using a multiethnic sample from the UK Biobank and All of Us cohorts. We specifically focus on examining the shared genetic architecture of cannabis use and liability to sleep problems as captured by formal diagnosis and medication prescription for sleep disorders. Our work expands the individual genetic associations to 39 loci, strong enrichment in the brain, and shows a robust connection between sleep problems and cannabis use.

## METHODS

### Cohorts

#### UK Biobank

The United Kingdom Biobank (UKB) is a biobank composed of more than 500,000 individuals recruited from 2006 to 2010 in the UK. Participants were between the ages of 37 and 73 at the time of recruitment, and were residents of the UK. The biobank data is composed of genotyped and imputed genetic data, lifestyle measures through questionnaires, and electronic health record data.

The biobank genotyped 488,477 participants and allows the data to be accessible to approved researchers; 49,950 individuals were genotyped using the Affymetrix (Thermo Fisher Scientific) UK BiLEVE Axiom Array, and 438,427 were genotyped using the closely related Affymetrix UK Biobank Axiom Array. From the arrays there is information on up to 825,927 SNPs and short indels, prioritizing variants involved in disease and ancestry-specific markers so that the data can be easily imputed. The genetic data was captured from blood samples taken during initial enrollment, and processed in 106 sequential batches, providing genotype calls on 812,428 variants across 489,212 participants.

Imputation was performed by the TOPMed Informatics Research Center using the genotype and sequencing data and the TOPMed R2 panel, and the resulting data is accessible to approved researchers through the UK Biobank Research Access Platform (UKB-RAP). For more information on how imputation was performed, as well as for more information on the cohorts contributing to the TOPMed imputation panel, please see Taliun, D., Harris, D.N., Kessler, M.D. et al^10^.

Ancestry classification of UK Biobank participants for the GWAS analysis followed the methods of the Pan-UKB Project using the Human Genome Diversity Project-1000 Genomes (HGDP-1KG) harmonized reference dataset^11 12^. The HGDP-1KG is a high-quality dataset of 4,094 whole genomes from labelled diverse continental populations. Principal components analysis (PCA) was first performed on unrelated (KING kinship coefficient < 0.125) individuals in the HGDP-1KG dataset after pruning variants (500kb window, r2 = 0.10) to extract the top 10 PCs of ancestry (Manichaikul et al., 2010; Patterson, Price, & Reich, 2006; Purcell et al., 2007). We then projected individuals from the UK Biobank onto the HGDP-1KG PC space and trained a random forest classifier given continental ancestry labels from the HGDP-1KG cohort to assign ancestry to UK Biobank individuals based on their top 10 PC scores using the Python package scikit-learn ^13^. The minimum random forest probability for assignment to a particular ancestry group was 0.5 and we completed 20 iterations of this model. An individual was assigned to a specific ancestry group and included in the genome-wide association study if 20/20 iterations assigned them to that specific ancestry, otherwise individuals were excluded from the GWAS as not of the desired ancestry or admixed. Finally, PCA was then completed within each ancestry of UKB individuals to extract the top 10 PCs of ancestry for inclusion in the GWAS as covariates to adjust for population stratification within each sub-population. Individuals in the GWAS analysis were restricted to those of European ancestry.

Phenotyping for cannabis use was defined using field 29104 (“Have you used cannabis (marijuana, grass, hash, ganja, blow, draw, skunk, weed, spliff, dope), even if it was a long time ago?”) from the online mental wellbeing questionnaire sent to participants, with individuals who responded to “No” encoded as controls, and individuals who responded with an answer containing “Yes” were encoded as a case.

#### Ethics statement UK Biobank

The North West Multi-centre Research Ethics Committee (MREC) has granted the Research Tissue Bank (RTB) approval for the UKB that covers the collection and distribution of data and samples (http://www.ukbiobank.ac.uk/ethics/). Our work was performed under the UKB application number 6818. All participants included in the conducted analyses have given a written consent to participate, and this work uses data provided by patients and collected by the NHS as part of their care and support.

#### All of Us

The All of Us Research Program is a United States-based longitudinal study aimed at improving precision medicine by collecting a variety of information on participants who reflect a diverse US population. The program integrates electronic health records, survey responses, and genetic data from over 400,000 participants and integrates them into a secure research platform.

The All of Us Research Program provides genotyping data for over 1 million variants utilizing the Illumina Infinium Global Diversity Array-8 (GDA-8). Markers were selected in order to optimize for a diverse cohort of different ethnic backgrounds. All array samples were QC’ed in a systematic fashion, with the current release (CDRv8) from the research program providing data on 447,278 participants with array data. All data was accessed within the All of Us Researcher Workbench, under Controlled Tier approval.

To run the genome-wide association study, the short-read whole genome sequencing data was utilized, which covers 414,830 participants. Following the standard QC performed by the All of Us Research Program, the ACAF bgen set was used as input, containing variants with a population-specific allele frequency greater than 1% or a population-specific allele count greater than 100. Please refer to the All of Us support hub for more information regarding the QC and utilization of the genetic data.

Phenotyping for cannabis use was defined using responses from the Lifestyle survey, specifically to the concept code 1585636. If individuals responded to question 1585636, “In your LIFETIME, which of the following substances have you ever used?”, with answer code 1585637, “Which Drugs Used: Marijuana Use”, they were encoded as a case; all other individuals who responded to the survey and did not have answer code 1585637 were encoded as a control. We used activity tracking data measured with Fitbit to estimate objective sleep duration in individuals with cannabis use adjusting for age, sex and body mass index.

#### Ethics statement All of Us

The All of Us Research Program is a nationwide longitudinal cohort study approved by centralized institutional review board (IRB) oversight coordinated by the U.S. National Institutes of Health. All participants provided written informed consent for the collection and use of genetic, lifestyle, and health data in biomedical research from a diverse U.S. population.

### Genome-wide Association Analyses

Genome-wide association analyses were performed in each cohort using REGENIE v4.0. In the UK Biobank analysis, logistic regression was performed with the following covariates: age (calculated using p34, p52, and p29203), sex, genotyping array, and 10 principal components accounting for within-EUR ancestry. In the All of Us analysis, logistic regression was performed using the following covariates: age (calculated from survey response date and date of birth), sex, and 10 principal components accounting for within-EUR ancestry.

### Meta-analysis

We performed fixed effects meta-analysis using METAL inverse variance weighted method^14^.

### Conditional and Joint Analysis for Independent Associations

We used GCTA’s conditional and joint analysis (COJO) to identify conditionally independent association signals from the GWAS summary statistics using a stepwise model selection procedure to jointly fit variants in approximate linkage disequilibrium, yielding a set of approximately independent lead SNPs for each locus. GCTA was performed with version 2.05beta, using the sbayes S option, so that the variance of SNP effects was related to minor allele frequency^15^. COJO was performed using GCTA version 1.94.1, and the LD reference panel was composed of 50,000 randomly selected unrelated respondents of the questionnaire cohort that were of European ancestry.

### Observed Heritability & Genetic Correlations

The observed heritability for cannabis usage and genetic correlations with external summary statistics were calculated using LD Score Regression v1.0.1. The summary statistics were munged into the proper format using the munge_sumstats.py tool provided by LDSC, and the variants were merged with the HapMap3 SNPs list to only include those within the LD reference panel.

### Pathway Analysis (GProfiler)

Pathway analysis was performed using the nearest protein coding gene annotations as input to the online GProfiler tool^16^.

### Partitioned heritability analysis

To test whether SNP heritability for cannabis use was enriched in specific tissue groups, partitioned heritability analysis was conducted using stratified LD score regression (S-LDSC). A multi-tissue chromatin annotation framework was applied to evaluate enrichment across broad tissue categories, while conditioning on the standard baseline model. Tissue-specific enrichment was quantified using the regression coefficient, enrichment estimate, and corresponding *p* value for each annotation group (PMID: 26414678; PMID: 29632380).

### Mendelian Randomization & SMR Portal

Mendelian Randomization was performed using the R package TwoSampleMR version 0.7.4^17^. For further followup using eQTL datasets, the online SMR Portal tool was used, uploading the meta-analysis summary statistics and utilizing the eQTL_eQTLGen dataset for Blood & Whole Blood and all of the Brain eQTL datasets^18^.

### PLACO

PLACO is a tool utilized for identifying pleiotropic loci between two traits of interest^19^. PLACO was used by sourcing the R script from the associated github, labeled PLACO_v0.1.1.R.

### SUSIE-COLOC

Colocalization analysis was performed using the coloc R package and its compatibility with the Sum of Single Effects (SuSiE) R package. Using loci identified from the PLACO analysis, regions to run susie on where defined as the HG38 LD block containing the PLACO tophit utilizing this file (https://github.com/jmacdon/LDblocks_GRCh38/blob/master/data/deCODE_EUR_LD_blocks.bed). Fine-mapping was first performed in each phenotype and then colocalized using the coloc.susie() function on the susie objects.

### Fitbit analysis

**f**We analyzed Fitbit-derived sleep data from 90,787 All of Us participants who responded to the cannabis use survey and had at least seven nights of wearable sleep recordings (29,973 cannabis users and 60,814 controls). All associations were adjusted for age, sex, BMI, number of recording nights of fitbit use, alcohol frequency, and PHQ-2 depression score.

## RESULTS

### GWAS of cannabis use discovers 39 loci

We performed genome-wide association analysis of cannabis use in the UK Biobank and All of Us cohorts. We discovered thirteen independent genome-wide significant genetic associations in the UK Biobank, ten associations in the All of Us cohort and 39 associations in the combined meta-analysis (**Figure 1, Figure S1A & S1B**). These associations recapitulate the previously reported associations with lifetime cannabis use including *CADM2* ^8^ and discover over 20 previously unreported genetic loci. Independent hits identified by conditional joint analysis are labeled in red (**Table 1**).

**Table 1.** Independent loci identified through conditional analysis (COJO).

| rsID | hg38 CHROM | hg38 POS | Nearest Protein Coding Gene | EA | NEA | EAF | Beta | SE | Pvalue | OR | OR_lower95 | OR_upper95 |
| --- | --- | --- | --- | --- | --- | --- | --- | --- | --- | --- | --- | --- |
| rs3927084 | 16 | 28627726 | SULT1A1 | A | C | 0.402 | -0.044 | 0.005 | 8.80E-17 | 0.957 | 0.947 | 0.967 |
| rs35344466 | 3 | 85583871 | CADM2 | A | C | 0.232 | 0.049 | 0.006 | 1.61E-15 | 1.050 | 1.038 | 1.063 |
| rs1788785 | 18 | 23562376 | NPC1 | T | C | 0.657 | 0.043 | 0.006 | 5.39E-15 | 1.044 | 1.033 | 1.055 |
| rs66632973 | 3 | 85536402 | CADM2 | A | T | 0.620 | -0.040 | 0.005 | 1.38E-13 | 0.960 | 0.950 | 0.971 |
| rs16867990 | 4 | 31128176 | PCDH7 | T | G | 0.294 | -0.040 | 0.006 | 1.37E-12 | 0.960 | 0.950 | 0.971 |
| rs62263112 | 3 | 117896668 | LSAMP | T | C | 0.121 | -0.056 | 0.008 | 1.96E-12 | 0.945 | 0.931 | 0.960 |
| rs2155290 | 11 | 112980346 | NCAM1 | C | G | 0.391 | 0.037 | 0.005 | 2.94E-12 | 1.038 | 1.027 | 1.049 |
| rs203456 | 17 | 19916911 | AKAP10 | C | T | 0.393 | -0.035 | 0.005 | 9.59E-11 | 0.966 | 0.955 | 0.976 |
| rs3101339 | 1 | 72282986 | NEGR1 | A | C | 0.658 | 0.040 | 0.006 | 1.53E-10 | 1.040 | 1.028 | 1.053 |
| rs4363528 | 10 | 102895758 | BORCS7-ASMT | A | G | 0.305 | 0.036 | 0.006 | 1.83E-10 | 1.036 | 1.025 | 1.048 |
| rs72813607 | 17 | 31134507 | NF1 | G | A | 0.130 | -0.049 | 0.008 | 1.92E-10 | 0.952 | 0.938 | 0.967 |
| rs2647239 | 4 | 105296192 | TET2 | A | G | 0.418 | -0.034 | 0.005 | 2.02E-10 | 0.967 | 0.957 | 0.977 |
| rs9297901 | 8 | 92024567 | RUNX1T1 | T | C | 0.232 | -0.038 | 0.006 | 1.63E-09 | 0.963 | 0.951 | 0.975 |
| rs10129426 | 14 | 103552118 | BAG5 | G | A | 0.540 | 0.031 | 0.005 | 1.76E-09 | 1.032 | 1.021 | 1.042 |
| rs6734603 | 2 | 181174002 | UBE2E3 | T | C | 0.737 | 0.035 | 0.006 | 1.98E-09 | 1.036 | 1.024 | 1.048 |
| rs9372607 | 6 | 97871615 | MMS22L | T | A | 0.881 | 0.048 | 0.008 | 3.11E-09 | 1.049 | 1.033 | 1.066 |
| rs35030637 | 1 | 41351014 | FOXO6 | C | T | 0.226 | -0.037 | 0.006 | 3.57E-09 | 0.964 | 0.952 | 0.976 |
| rs40465 | 5 | 104646025 | NUDT12 | T | G | 0.334 | 0.032 | 0.006 | 4.29E-09 | 1.033 | 1.022 | 1.044 |
| rs4925119 | 17 | 17839590 | SREBF1 | A | G | 0.641 | 0.032 | 0.005 | 4.80E-09 | 1.032 | 1.021 | 1.043 |
| rs1854465 | 10 | 81436153 | NRG3 | C | T | 0.854 | -0.043 | 0.007 | 4.88E-09 | 0.958 | 0.944 | 0.972 |
| rs115882849 | 3 | 54122697 | CACNA2D3 | G | A | 0.149 | 0.042 | 0.007 | 6.32E-09 | 1.043 | 1.028 | 1.058 |
| rs68153678 | 3 | 147486033 | ZIC1 | A | G | 0.365 | -0.031 | 0.005 | 6.79E-09 | 0.969 | 0.959 | 0.979 |
| rs10865392 | 2 | 72574624 | EXOC6B | A | C | 0.884 | -0.047 | 0.008 | 1.04E-08 | 0.954 | 0.939 | 0.970 |
| rs34324193 | 18 | 46469087 | RNF165 | T | C | 0.330 | -0.031 | 0.006 | 1.27E-08 | 0.969 | 0.959 | 0.980 |
| rs10228057 | 7 | 1840143 | MAD1L1 | A | T | 0.379 | -0.031 | 0.005 | 1.31E-08 | 0.970 | 0.960 | 0.980 |
| rs4658588 | 1 | 243682969 | AKT3 | A | G | 0.689 | 0.031 | 0.006 | 2.06E-08 | 1.032 | 1.021 | 1.043 |
| rs10863714 | 1 | 208537248 | PLXNA2 | A | G | 0.570 | -0.029 | 0.005 | 2.19E-08 | 0.971 | 0.961 | 0.981 |
| rs13107325 | 4 | 102267552 | SLC39A8 | C | T | 0.076 | -0.054 | 0.010 | 3.17E-08 | 0.947 | 0.929 | 0.966 |
| rs10215276 | 7 | 127869390 | SND1 | C | T | 0.879 | -0.044 | 0.008 | 3.18E-08 | 0.957 | 0.943 | 0.972 |
| rs4766578 | 12 | 111466567 | ATXN2 | T | A | 0.498 | -0.029 | 0.005 | 3.80E-08 | 0.972 | 0.962 | 0.982 |
| rs62380518 | 5 | 63689240 | HTR1A | G | A | 0.133 | 0.042 | 0.008 | 3.95E-08 | 1.043 | 1.028 | 1.059 |
| rs6739515 | 2 | 125266661 | CNTNAP5 | G | A | 0.275 | -0.032 | 0.006 | 4.19E-08 | 0.969 | 0.958 | 0.980 |
| rs2209574 | 13 | 71091627 | DACH1 | G | A | 0.604 | -0.029 | 0.005 | 4.46E-08 | 0.971 | 0.961 | 0.982 |
| rs11765366 | 7 | 101153587 | AP1S1 | C | G | 0.198 | -0.036 | 0.007 | 4.51E-08 | 0.965 | 0.952 | 0.977 |
| rs73011961 | 2 | 147647337 | ACVR2A | C | A | 0.103 | -0.046 | 0.009 | 4.80E-08 | 0.955 | 0.939 | 0.971 |
| rs57617625 | 2 | 80169139 | CTNNA2 | C | G | 0.328 | -0.031 | 0.006 | 4.87E-08 | 0.970 | 0.959 | 0.981 |

**Figure 1.**
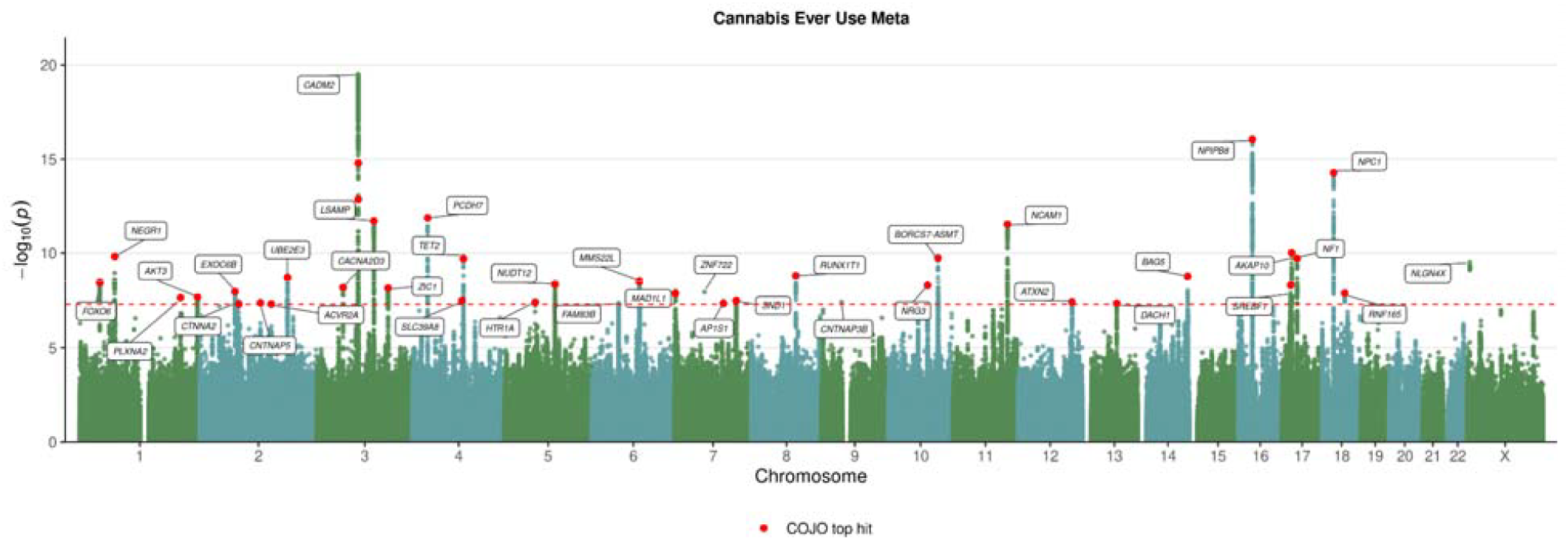
Manhattan Plot of META analysis of cannabis use in European ancestry.

### Genetic variants for cannabis use are enriched in brain tissues

We observe associations with neuronal genes including serotonin receptor 1A (*HTR1A*), calcium channel genes (including *CACNA2D3*) and additionally several well-established neuronal genes including *NF1, NPC1* and *NCAM1* supporting a role of neuronal function in cannabis use. Additionally, we observed several associations at canonical sleep and circadian genes. These included *CADM2* which in addition to associating with cannabis use shows genome-wide association with clinical insomnia^20^, daytime napping and sleepiness^21^ and modulating hypersomnia in model organisms^22^. In addition, we saw associations with *HTR1A*, a target for REM sleep suppressing 5-HT1A receptor agonist SEP-363856^23^, MAD1L1 that has been previously associated with sleep duration^24^, *RAI1* is associated with chronotype and circadian preference and *SLC39A8* that has been associated with multiple sleep^25^ and neuropsychiatric traits ^26^. These neuronal and sleep associations prompted us to formally test the tissue specific enrichment of genetic architecture of cannabis use. We observed that even when using all tissues from GTEx the tissue enrichment was only enriched in the brain (**Figure 2a, Table S1**). This finding was supported by stratified LDSC that showed the strongest enrichment in the CNS and brain cell types showing the strongest enrichment (**Figure 2b,d, Table S2,3**), and pathway analysis that had a significant enrichment for brain morphogenesis and FOXO-mediated transcription (**Table S4**). Additionally, we performed summary-based Mendelian Randomization (SMR) using SMR Portal and found 10 independent lead variants with significant evidence for colocalization between cannabis use and Brain QTL datasets (**Figure 2c, Table S5**).

**Figure 2.**
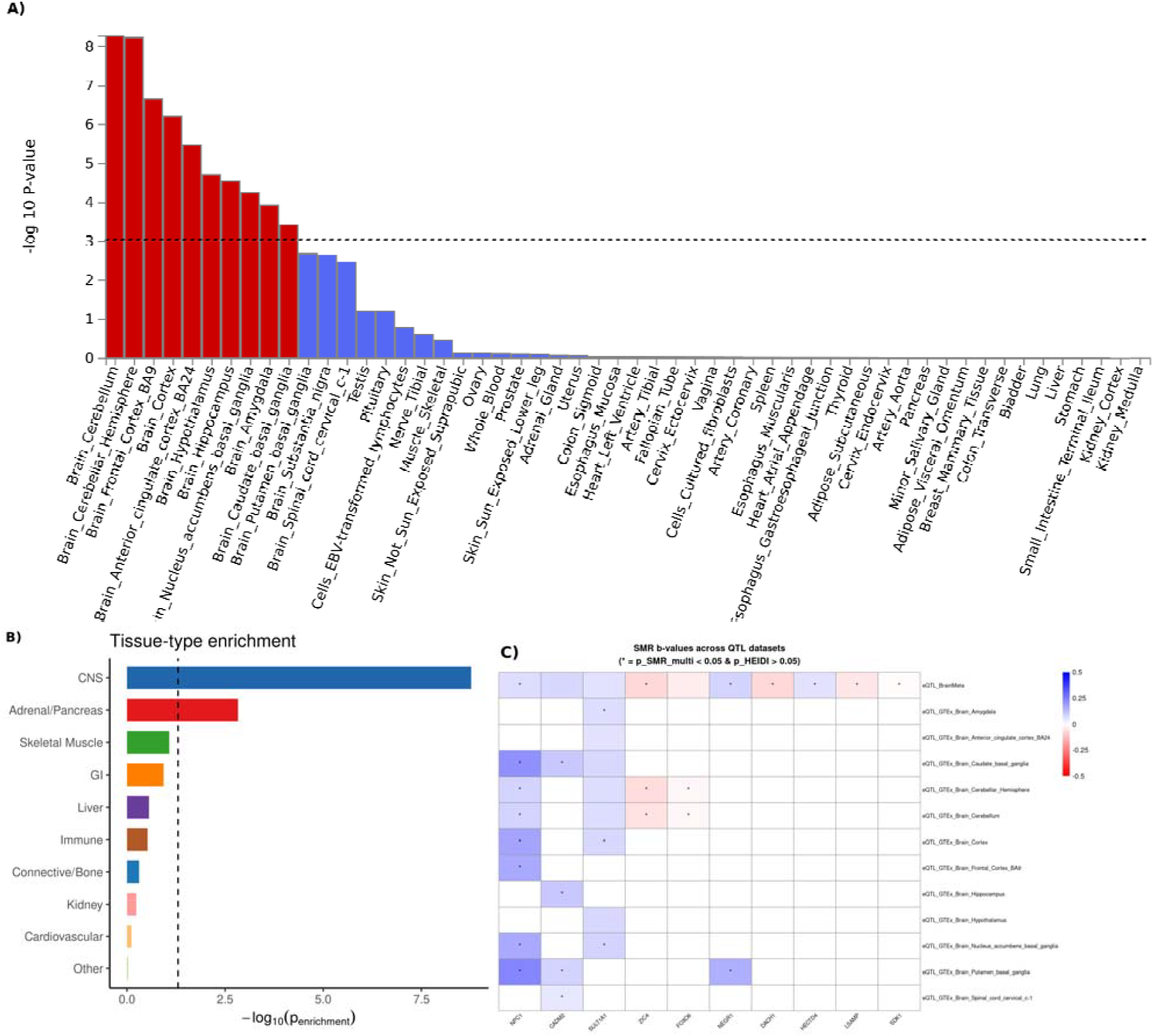
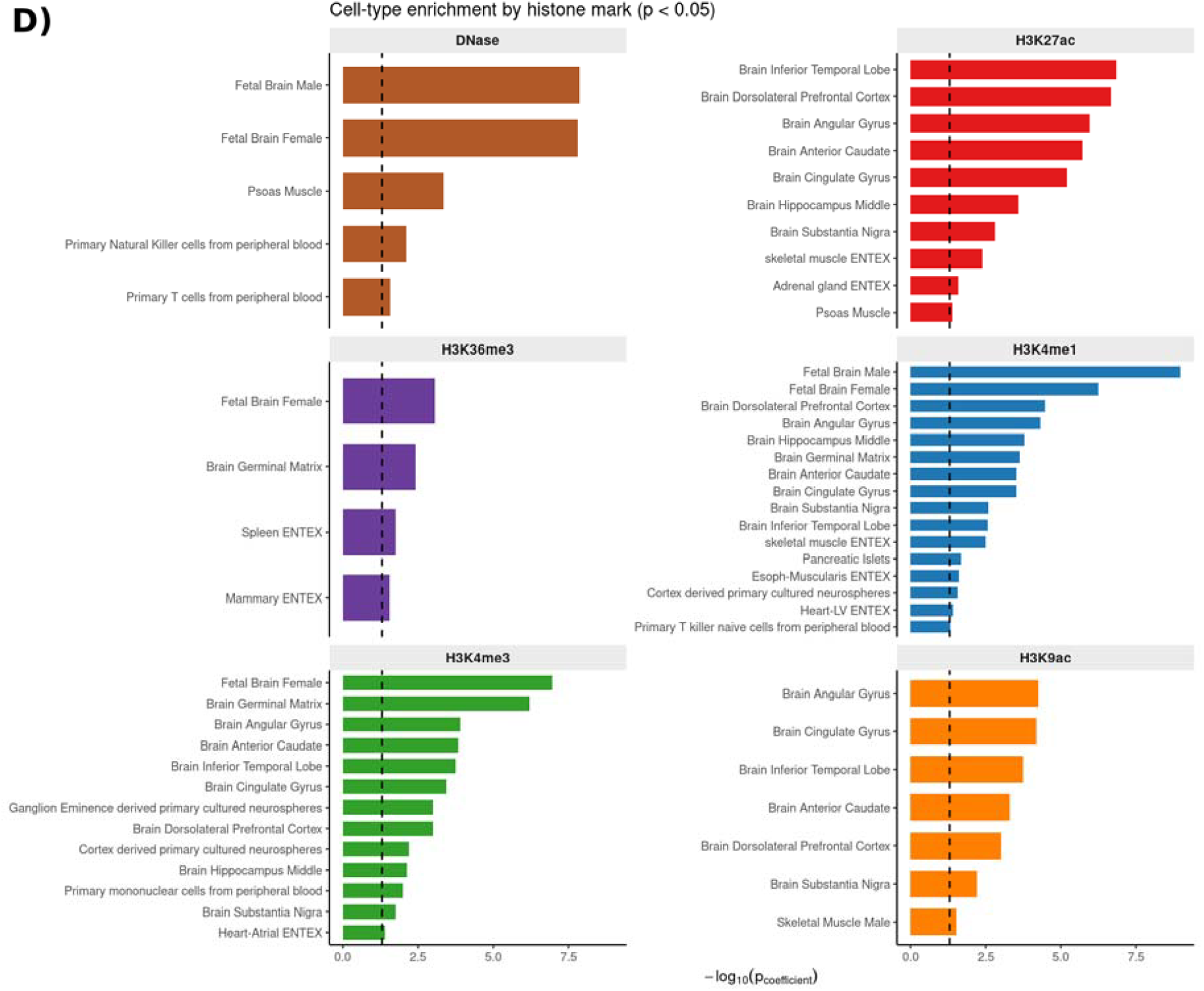
Enrichment of cannabis use summary data primarily seen in brain tissue types and brain cell types. **A)** Tissue Enrichment analysis in FUMA showing enrichment isolated to the brain. **B)** Stratified LDSC analysis by tissue, demonstrating the strongest enrichment within the central nervous system (CNS). **C)** Tile plot of Summary-based Mendelian Randomization (SMR) analysis showing significant colocalization between cannabis use and Brain QTL datasets across 10 independent lead variants. **D)** Stratified LDSC analysis highlighting enrichment by specific brain cell types

### Cannabis use shares genetic architecture with sleep and circadian traits

Sleep problems are one of the most common reasons for cannabis use. To better understand the relationship between sleep problems and cannabis use we computed genetic correlation between cannabis use and clinical sleep disorders, habitual sleep traits, and circadian preference traits. Of the circadian preference traits, we observed negative correlations with morningness chronotype (rg = -0.198, P = 3 x 10^-19^), as well as a negative correlation with ease of awakening (rg = -0.194, P = 1 x 10^-14^) (**Table S6**). We also observed that cannabis use was positively correlated with insomnia (rg = 0.184, P = 4 x 10^-10^) and with insomnia medications (rg = 0.286, P =1 x 10^-24^) (**Figure 3a**). To further contextualize the genetic effects of sleep within daily rhythms, we then performed genetic correlations with total log acceleration for physical activity in 2 hour binned windows across the day (**Figure 3b, Table S7**). This resulted in a clear positive relationship between the genetics of cannabis use in the context of sleep, whereas there was a negative correlation with daytime activity measurements and positive correlation with increased activity at night. Consistently, using activity tracking data from All of Us, we observe longer sleep duration in individuals who use cannabis (sleep duration cases = 7.10 h, sleep duration controls = 7.006 h, P = 1.10e-18).

**Figure 3.**
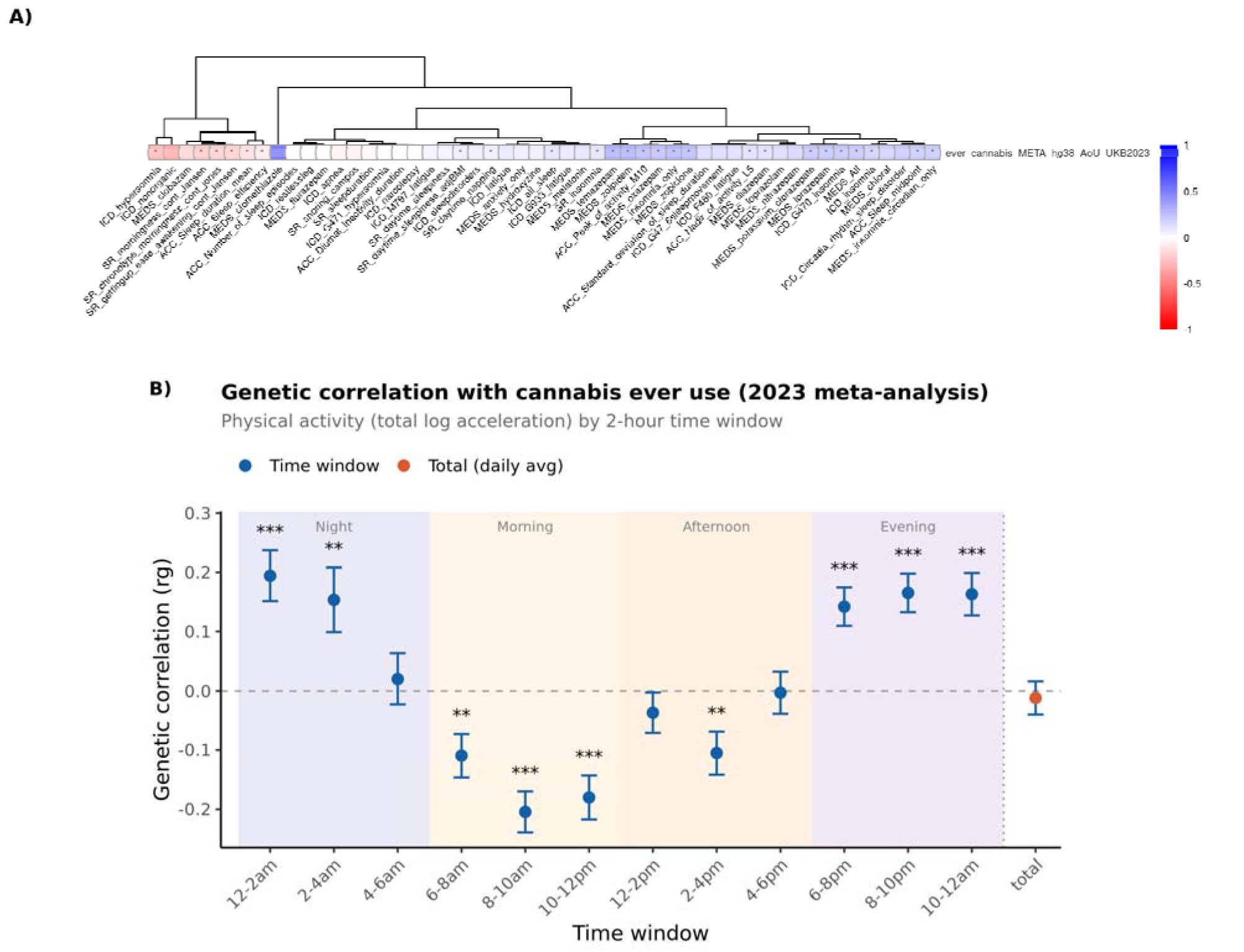
Genetic correlation between cannabis use and sleep traits, with finemapping followup using PLACO and SuSiE Coloc. **A)** Genetic correlation heatmap of cannabis use with diagnostic code and self-report sleep phenotypes, accelerometry, and medication-based phenotypes, illustrating positive correlations with insomnia and insomnia medications. **B)** Genetic correlations with total log acceleration for physical activity analyzed in 2-hour binned windows across the day.

In addition, using PLACO to identify plausible shared regions of genetic signal, and SUSIE-COLOC we examined shared genetic signals between clinical insomnia and cannabis use (**Figure S2**). We identified a near-perfect colocalization at chromosome 4 SLC39A8 with posterior probability = 1 between cannabis use and clinical insomnia. Additionally, NCAM1 and CADM2 loci both co-localized between clinical insomnia and cannabis use (pp.H4 > 0.7) further supporting a joint genetic architecture at single locus level between sleep and cannabis use in key cannabis loci (**Table S8**).

This correlation was additionally supported by Mendelian randomization where we saw a causal role from insomnia to cannabis use (P = 0.00566). Furthermore, cannabis use associated with increased diagnosis of insomnia in MR (P = 0.02) (**Table S9**). These findings align with the established literature that connects cannabis use with sleeping problems.

### Cannabis use with REM and NREM phenotypes

One of the best established phenotypic associations with cannabis use is the effect on dreaming where cannabis use suppresses REM sleep. The majority of dreams occur in REM sleep and cannabis use consequently is associated with dreamless sleep. However, stopping cannabis use causes REM rebound where REM sleep dominates during the night and causes intense and sometimes scary dreams and nightmares. To study the genetic overlap between cannabis use and REM sleep, we utilized recently published summary statistics on REM and non-REM (NREM) sleep.

## DISCUSSION

Here we examined cannabis use in 370,000 individuals with information on cannabis use from UK Biobank and All of Us cohorts. Our analysis expands the known associations to 39 independent loci, discovers a strong neuronal component with cannabis use, and highlights a strong shared genetic architecture between cannabis use and sleep traits.

In this study, we discovered a robust association with canonical neurotransmitter and synaptic genes, many of which have been implicated in neuropsychiatric and sleep regulation. In particular, we find an association with CADM2 that faithfully replicates across different cohorts and settings when examining cannabis use ^8,27^. CADM2 has been previously associated with clinical insomnia^20^. It is part of the synaptic cell adhesion molecule 1 (SynCAM) family and similar other canonical neuronal genes are also reflected in the meta-analysis.

The meta-analysis also yielded significant findings in three key proteins related to memory formation and activity, supporting the hypothesis of cannabis use being associated with memory impairment. NCAM1, a neural cell adhesion molecule widely involved in neuronal activity and also a member of the Immunoglobulin (Ig) superfamily was found to be significant, and has been associated with long term memory formation. FOXO6 was also found to be associated in the meta-analysis, with its expression being specific to the central nervous system. Knockout of FOXO6 in mice has been noted to lead to memory consolidation defects^28^. This is particularly interesting as another significant gene, AKT3, is involved in the phosphorylation and inactivation of FoxO transcription factors, potentially hinting at a potential biological pathway relevant to cannabis use and impaired memory. Overall, this study supports the association of a broad neuronal set of regulators in cannabis use.

NPC1 was also identified through conditional analysis of the meta-analysis, potentially elucidating another mechanism involved in the consumption of cannabis. Cannabis is known to be highly lipophilic, potentially affecting lipid rafts or non-lipid rafts within the cell membrane where receptors like CB1 is located, and thus disrupting the endocannabinoid system^29,30^. This is interesting, especially within the context of higher percentage cannabis products, as a flood of highly lipophilic molecules could affect the machinery involved with the endocannabinoid system and lead to accumulation in other components, such as the lysosome where NPC1 is primarily located. Variation in the NPC1 gene is still actively studied, with current research noting NPC1 has been involved with lysosomal storage complications and progressive neurodegeneration in the context of Niemann-Pick disease type C (NPC). Also, individuals with NPC have been shown to have hypocretin (orexin) neuron loss, mirroring symptomatology of narcolepsy type 1 and cataplexy symptoms^31^. SMR provides evidence for colocalization between NPC1 eQTL data and cannabis use across eight phenotypes, the most out of all genes identified through this analysis (**Figure 2b**). This provides an interesting link between lysosomal storage functionality and sleep/wake circuitry particularly in the context of cannabis use, but more research is needed to fully understand the context of this association.

The genetic architecture of cannabis use was essentially fully enriched in the brain tissue. We observed this at the level of FUMA brain tissue enrichment, stratified LDSC and single locus associations. These findings are in line with earlier reports of cannabis use, and with cannabis use disorder that have shown several neuronal associations including the canonical CADM2 gene. Our data together with the earlier reported association expands the set of neuronal molecules that are associated with cannabis use and connect them with sleep regulation.

One of the most exciting findings in the current study was the association with individual genes that are known regulators of sleep and circadian rhythms. We identify associations with *HTR1A, MAD1L1* and *SLC39A8* that all have been associated with sleep traits. In addition, *HTR1A* agonists affect REM sleep^32^, which aligns with the well-established role of cannabis suppressing REM sleep amount.

In this study, we additionally observed a positive genetic correlation between sleep problems and cannabis use. Earlier GWAS on cannabis use did not find a strong genetic correlation with insomnia symptoms. The differences between the studies are potentially driven by the earlier study focusing on self-reported symptoms of insomnia from earlier questionnaire data, whereas the current study included clinical diagnosis of insomnia and insomnia medication use, and self-reported data from the UK Biobank baseline and updated sleep questionnaires. Overall, the current findings support the connection between cannabis use and sleep problems, particularly insomnia and delayed circadian preference.

Finally, we discovered a significant genetic correlation between cannabis use and dreaming. Earlier epidemiological and clinical studies on cannabis have systematically reported the strong effect of cannabis on REM sleep where cannabis use first suppresses dreaming and after stopping cannabis use, causes REM rebound and vivid dreams. In our study cannabis use was associated with higher dream recall and higher frequency of nightmares. This finding is intriguing and has several possible mechanisms. For example, nightmares and dream recall are heavily influenced by insomnia with higher insomnia increasing dream recall and nightmares^33^at population setting and at sleep laboratory studies^34^. The association between cannabis use and dreaming may be modified by insomnia. Additionally, individuals who use cannabis will experience at least one REM rebound that typically causes intense and vivid dreams including nightmares. The use of cannabis can counterintuitively increase REM rebound and dreaming. Since our data is with a one timepoint questionnaire for cannabis use, we cannot fully resolve these two possible mechanisms in the current data. Finally, it is possible that individuals who experience vivid dreams, or have liability for nightmares use cannabis as a REM suppressant. This hypothesis would need to be examined in additional studies.

Overall, we show that cannabis use is a robust genetic trait. Its genetic architecture is primarily localized in brain tissues, with several canonical neuronal genes associating with cannabis use. In the current work we discover that sleep and circadian preference are strongly associated with cannabis use with insomnia, delayed circadian preference and dreaming having a key role in cannabis use. Future work is still required in order to fully understand the effects of cannabis on sleep regulation and recovery, especially the consideration of different strains of cannabis and their effects on biological processes. Overall these findings highlight the complex genetic architecture of cannabis use and its emerging shared genetic mechanisms in connection with sleep.

## Data Availability

All data produced in the present study are available upon reasonable request to the authors

